# A Retrospective Analysis of Feeding Practices and Complications in Patients with Critical Bronchiolitis on Non-invasive Respiratory Support

**DOI:** 10.1101/2021.04.15.21255583

**Authors:** Ariann Lenihan, Vannessa Ramos, Nichole Nemec, Joseph Lukowski, Junghyae Lee, K. Meghan Kendall, Sidharth Mahapatra

## Abstract

Limited data exist regarding feeding pediatric patients managed on non-invasive respiratory support (NRS) modes that augment oxygentation and ventilation in the setting of respiratory failure. We conducted a retrospective cohort study to explore the safety of feeding patients managed on NRS with acute respiratory failure secondary to bronchiolitis. Children up to 2 years old with critical bronchiolitis managed on RAM, CPAP, or BiPAP were included. Of the 178 eligible patients, 64 were reportedly NPO while 114 were fed (EN). Overall equivalent in severity of illness, younger patients populated the EN group, while the NPO group experienced a higher incidence of intubation. Duration of PICU stay and NRS were shorter in the NPO group, though intubation eliminated the former difference. Within the EN group, ninety percent had feeds initiated within 48 hours and 94% reached full feeds within 7 days of NRS initiation, with an 8% complication and <1% aspiration rate. Reported complications did not result in escalation of respiratory support. Notably, a significant improvement in heart rate and respiratory rate was noted after feeds initiation. Taken together, our study supports the practice of early enteral nutrition in patients with critical bronchiolitis requiring non-invasive respiratory support.

## 1. Introduction

Viral bronchiolitis is the most common lower respiratory tract illness and a leading cause of hospitalization of infants and young children.[1-3] Between 2-25% of admitted bronchiolitis patients will suffer acute respiratory failure, necessitating admission to the pediatric intensive care unit (PICU).[4-6] Current mainstay of therapy emphasizes supportive care.[3] Non-invasive respiratory support (NRS) refers to the delivery of oxygen and provision of respiratory support through modalities that can allay the need for endotracheal intubation. These include continuous positive airway pressure (CPAP), bilevel positive airway pressure (BiPAP), and RAM cannula, which is a modified BiPAP delivery system that employs nasal cannula and is well tolerated by children under 2 years of age. NRS has been studied as first line treatment for acute respiratory failure and shown to be well tolerated, and has become the preferred mode of treating acute hypoxic and hypercarbic respiratory failure secondary to bronchiolitis.[7-10]

Despite widespread use of enteral and parenteral nutrition in critically ill children, caloric and protein underfeeding continue to remain a common problem, including amongst the viral bronchiolitis population.[11, 12] Previous studies have revealed NRS as a common factor for both delayed EN initiation and underfeeding in the PICU.[13, 14] Despite data to the contrary, providers often cite safety concerns for delaying EN on NRS.[15, 16] Commonly cited reasons include: a) the potential for the patient’s status to worsen potentially requiring mechanical ventilation; b) nasogastric tubes interfering with optimal NRS mask seal; c) exacerbating respiratory failure from breaks to allow oral feeds; and d) concerns surrounding aspiration of gastric contents during feeds.[17, 18] Notably, pediatric patients often require sedation to tolerate NRS, which further raises the concern for aspiration from relaxation of airway protective reflexes.[19, 20] Overall, there lacks consensus on the risk-benefit of feeding patients on NRS.

In this study, we aimed to examine the safety of enterally feeding pediatric patients with acute respiratory failure due to viral bronchiolitis managed on NRS by retrospectively examining the feeding practices within our PICU and assessing complications associated with enteral feeding. We also aimed to understand the possible benefits of early enteral nutrition on physiometric parameters and sedation needs.

## 2. Materials and Methods

### Setting

This study was conducted at the Children’s Hospital and Medical Center, Omaha, which is the only free-standing pediatric hospital in the state of Nebraska. Affiliated with the University of Nebraska Medical Center, this 145-bed tertiary pediatric medical center currently houses a 27-bed combined cardiac/non-cardiac pediatric intensive care unit with an annual admission rate of ∼1100 patients, an average daily census of 20.4, and a Standardized Mortality Ratio of 0.71.

### Study Design

After obtaining approval from our institutional review board, we performed a retrospective chart review of all pediatric patients with critical bronchiolitis admitted to our PICU between January 2015 and December 2017. Informed consent was waived given the minimal risk nature of this study. This study adhered to the ethical principles outlined in the Declaration of Helsinki as amended in 2013 and was HIPAA compliant.

### Eligibility

Eligible patients were: 1) ≥37 weeks corrected gestational age, older than 72 hours and ≤2 years old at admission, 2) carrying an ICD diagnosis of acute bronchiolitis, and 3) managed on non-invasive respiratory support for acute respiratory failure, including RAM cannula, CPAP and/or BiPAP. Exclusion criteria included: 1) either needing intubation within the first 24 hours of admission to PICU or never requiring NRS during PICU stay, 2) chronic ventilator dependence, 3) immediate postoperative status, 4) single ventricle physiology, 5) active gastrointestinal bleed, 6) short-gut syndrome, 7) chronic TPN dependence, and 8) any do-not-resuscitate (DNR) or other limitations in care.

### Variables

Demographic and clinical data collected included gender, weight, gestational age, history of prematurity, underlying neurologic or genetic abnormalities, dates of admission and discharge from PICU, disease severity scores, mortality, presence and type of infecting pathogen. Respiratory support data collected included type and duration of NRS, incidence and duration of intubation, extubation success, and ventilator-free-days. Nutrition data collected included mode and route of nutrition, time to initiation and to full nutrition on NRS, reported complications, and any evidence of aspiration. To evaluate the effect of enteral nutrition on comfort and sedation needs, data on sedation use while on NRS (type, duration, dosage) and physiometric data (heart rate and respiratory rate) prior to and after feeding initiation were collected.

### Definitions

Critical bronchiolitis was defined as viral bronchiolitis leading to acute respiratory failure requiring admission and management in a pediatric intensive care unit with high risk for adverse outcomes. Acute respiratory failure was diagnosed in patients requiring non-invasive respiratory support to mitigate work of breathing and/or to keep oxygen saturation >88%. Types of non-invasive respiratory support included in this study were CPAP, BiPAP, or RAM cannula; although heat high-flow nasal cannula (HHFNC) is a form of non-invasive respiratory support, authors did not include this mode because most patients on HHFNC are managed on the pediatric floor at our institution and would not satisfy the definition of critical bronchiolitis. Our unit’s practice for utilization of RAM cannula is to provide a set PIP, PEEP, i-time and respiratory rate utilizing a Servo ventilators. CPAP, defined as continuous positive airway pressure, was provided via nasal or face-mask through a Servo ventilator; BiPAP, defined as bilevel positive airway pressure, was similarly provided through nasal or face-mask through a Servo ventilator. Ventilator-free-days were defined as described previously.[21] Extubation failure was defined as re-intubation within 24 hours of liberation from invasive positive pressure ventilation. Any patient receiving enteral nutrition, by oral, nasogastric, nasojejunal, gastrostomy or jejunostomy tubes, while on NRS, was assigned to the enteral nutrition (EN)group; patients who did not receive any feeds while on NRS were assigned to the NPO group. Optimal time to feeding initiation (within 48 hours) and to full feeds (within 7 days) were defined based on the American Society for Parenteral and Enteral Nutrition (ASPEN).[22] Complications after feeding initiation were recorded and are detailed in **Supplementary Table 3**. Aspiration was defined as any documented feeds found in the nasopharynx concurrent with increased work of breathing. Physiometric parameters refer to heart rate and respiratory rate that were recorded for the EN group prior to and after feeds initiation.

### Statistical Analyses

For two group comparisons between the NPO and EN groups, the Wilcoxon rank-sum test or the chi-square test of independence was used. Continuous variables are presented with medians and interquartile ranges, while categorical variables are described using frequencies and percentages. Demographic and clinical data relating to age, NRS duration, PICU length of stay, pathogen burden, intubation duration, and ventilatory-free-days were compared between groups using the Wilcoxon rank-sum test. All other data were compared using the chi-square test of independence. Nutrition data for the EN group is presented as frequencies and percentages. Physiometric parameters (heart rate and respiratory rate) and sedation use between groups were compared using the Mann-Whitney U test. Statistical significance was established at *p* <0.05.

## 3. Results

### 3.1 Eligibility

We identified 342 pediatric patients admitted to our pediatric intensive care unit between January 2015 and December 2017 with a diagnosis of critical bronchiolitis. After excluding 164 patients, for never requiring NRS (75), requiring intubation within the first 24 hours (65), having chronic ventilator dependence (12), falling out of the age criteria (6), having a single-ventricle physiology (4), or being fresh post-operative status (2), 178 eligible critically-ill pediatric patients were stratified into nil per os (NPO, *n*=64) and enteral nutrition (EN, *n*=114) groups based on their feeding status during their critical illness (**Figure 1**).

**Figure 1.**
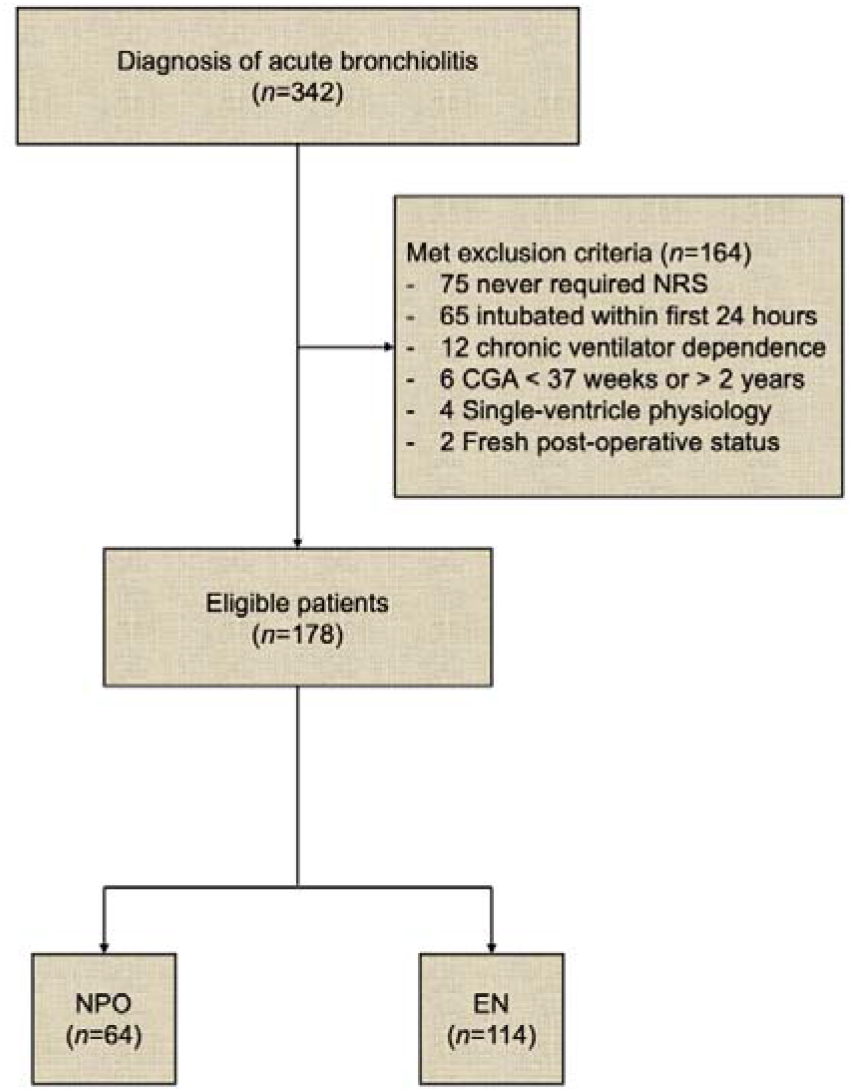
Flow diagram of patient enrollment. CGA = corrected gestational age, PICU = pediatric intensive care unit, CPAP = continuous positive airway pressure, BiPAP = bilevel positive airway pressure, NRS = non-invasive respiratory support, NPO = nil per os (nothing by mouth), EN = enteral nutrition.

### 3.2 Patient Demographics

When comparing demographic and clinical data between the two groups, there were no significant differences in gender, past medical history of prematurity, underlying genetic or neurologic abnormalities, severity of illness, or overall mortality. However, median age in the EN group was significantly lower than that of the NPO group (3 months, IQR 2-6 vs. 10 months, IQR 3-16; *p* <0.001) with a higher proportion of infants in the former group (82% vs. 48%; *p* <0.001). Not surprisingly, median weight was also lower in this group (5.7 kg, IQR 4.7-7.4 vs. 8.4 kg, IQR 5.4-10.5; *p* <0.001) (**Table 1**).

**Table 1.**
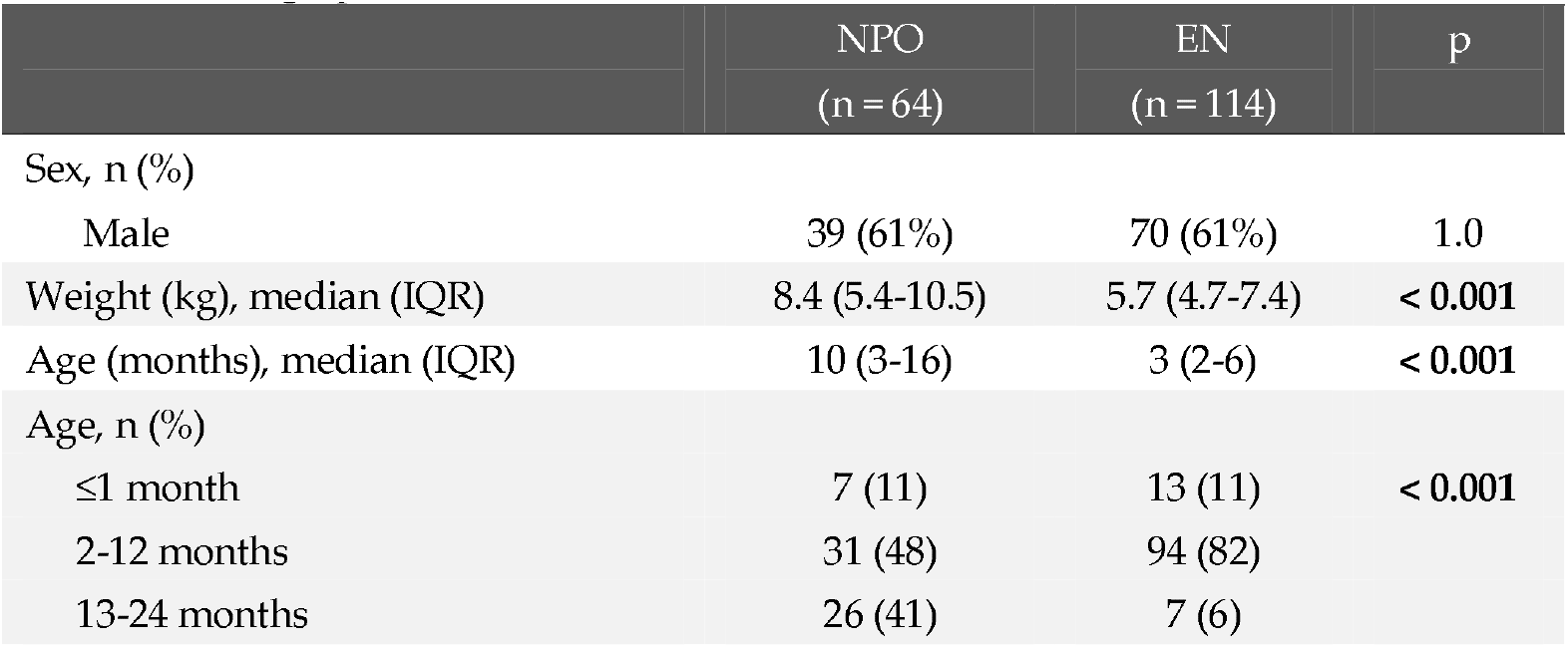
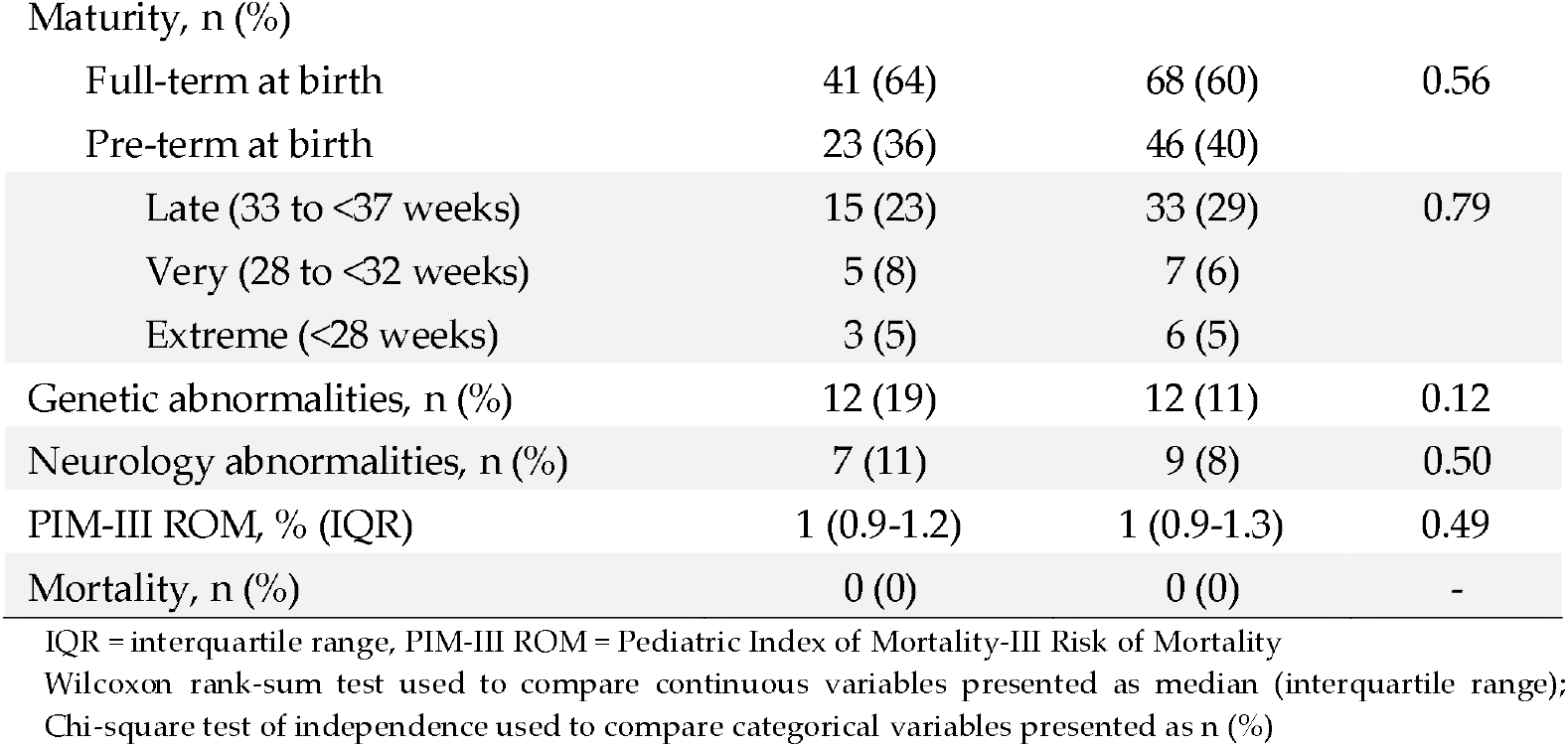
Demographic Data for Entire Cohort (n = 178)

|  | NPO<br>(n = 64) | EN<br>(n = 114) | p |
| --- | --- | --- | --- |
| Sex, n (%) |  |  |  |
| Male | 39 (61%) | 70 (61%) | 1.0 |
| Weight (kg), median (IQR) | 8.4 (5.4-10.5) | 5.7 (4.7-7.4) | < 0.001 |
| Age (months), median (IQR) | 10 (3-16) | 3 (2-6) | < 0.001 |
| Age, n (%) |  |  |  |
| ≤1 month | 7 (11) | 13 (11) | < 0.001 |
| 2-12 months | 31 (48) | 94 (82) |  |
| 13-24 months | 26 (41) | 7 (6) |  |
| Maturity, n (%) |  |  |  |
| Full-term at birth | 41 (64) | 68 (60) | 0.56 |
| Pre-term at birth | 23 (36) | 46 (40) |  |
| Late (33 to <37 weeks) | 15 (23) | 33 (29) | 0.79 |
| Very (28 to <32 weeks) | 5 (8) | 7 (6) |  |
| Extreme (<28 weeks) | 3 (5) | 6 (5) |  |
| Genetic abnormalities, n (%) | 12 (19) | 12 (11) | 0.12 |
| Neurology abnormalities, n (%) | 7 (11) | 9 (8) | 0.50 |
| PIM-III ROM, % (IQR) | 1 (0.9-1.2) | 1 (0.9-1.3) | 0.49 |
| Mortality, n (%) | 0 (0) | 0 (0) | - |

IQR = interquartile range, PIM-III ROM = Pediatric Index of Mortality-III Risk of Mortality
Wilcoxon rank-sum test used to compare continuous variables presented as median (interquartile range);
Chi-square test of independence used to compare categorical variables presented as n (%)

### 3.3 Clinical Characteristics

When examining ventilation and pathogen characteristics for all patients, groups were similar with respect to the type of NRS (RAM vs. CPAP vs. BiPAP), pathogen number and type. More specifically, a majority of patients in both groups were managed on RAM cannula (≥95%); had a single pathogen etiology (>70%) causing critical bronchiolitis, and mostly viral in origin (>90%) **(Table 2, Table S2)**. However, the NPO group had both shorter median NRS duration (1 day, IQR 0.84-2 vs. 3 days, IQR 2-4; *p*<0.001) and overall PICU length of stay compared to the EN group (2 days, IQR 1-3 vs. 3 days, IQR 2-5; *p*<0.001) (Table 2); the latter difference was lost upon intubation (11 days, IQR 6-32.5 vs. 15 days, IQR 12.5-25.5; *p*=0.38, **Table S1**). Moreover, while intubation rates were generally low in the entire cohort, the NPO group had a 3.5-fold higher incidence of intubation compared to the EN group (14% vs. 4%; p=0.016) **(Table 2)**. Within this cohort of intubated patients, characterized by a high incidence of multi-microbial infections (median 3 pathogens per patient), the EN group had a significantly higher incidence of bacterial superinfection compared to the NPO group (100% vs. 45%), while the NPO group had a significantly shorter median NRS duration prior to intubation (1.5, IQR 1.1-2.3 vs. 2.5, IQR 1.9-4.5; *p*=0.044). No differences were discerned with respect to median weight, age, time to and duration of intubation, extubation failure, or ventilator-free-days. (**Table S1**).

**Table 2.** Clinical Characteristics of Entire Cohort (n = 178)

|  | NPO<br>(n = 64) | EN<br>(n = 114) | p |
| --- | --- | --- | --- |
| NRS duration (days), median (IQR) | 1 (0.84-2) | 3 (2-4) | < 0.001 |
| Type of NRS, n (%) |  |  |  |
| RAM | 52 (95) | 108 (99) | 0.34 |
| CPAP | 0 (0) | 1 (1) |  |
| BiPAP | 3 (5) | 2 (2) |  |
| Intubation<sup>#</sup>, n (%) | 9 (14) | 5 (4) | 0.016 |
| Pathogen<sup>f</sup> (#), median (IQR) | 1 (1-2) | 1 (1-1) | 0.21 |
| Pathogen positive, n (%) | 57 (89) | 105 (92) | 0.5 |
| Single | 41 (72) | 81 (77) | 0.53 |
| Multiple | 16 (28) | 25 (24) |  |
| Virus only | 53 (93) | 99 (94) | 0.74 |
| Virus + Bacteria | 4 (7) | 6 (6) |  |
| PICU LOS (days), median (IQR) | 2 (1-3) | 3 (2-5) | < 0.001 |

<sup>#</sup> Refer to Table S1 for details
<sup>f</sup> Refer to Table S2 for details
NRS = non-invasive respiratory support, IQR = interquartile range, CPAP = continuous positive airway pressure, BiPAP = bilevel positive airway pressure, PICU LOS = pediatric intensive care unit length of stay,
Wilcoxon rank-sum test used to compare continuous variables presented as median (interquartile range); Chi-square test of independence used to compare categorical variables presented as *n* (%)

### 3.4 Enteral Nutrition Details

Amongst the non-intubated EN group of patients (*n*=109), we delved deeper to dissect feeding practices and complications (**Table 3**). Overall, the mode and route of highest physician preference was continuous (78%) via the nasogastric route (63%). Ninety percent of patients had feeds initiated within 48 hours of NRS initiation (which corresponded closely with PICU admission) and 94% reached full caloric goal within 7 days. Median time to initiation of enteral feeds was 19 hours (IQR 11-37) and median time to full feeds was 40 hours (IQR 24-58) after NRS initiation. Complications with enteral nutrition were encountered in only 8% of patients with only 1 patient having documented evidence of aspiration (**Table S3**). Of note, none of these patients experienced any escalation in NRS support.

**Table 3.** Nutrition Details for Enteral Group (n = 109)

|  | n | % |
| --- | --- | --- |
| Route |  |  |
| PO | 11 | 10 |
| NG | 69 | 63 |
| NJ | 14 | 13 |
| GT/JT | 15 | 14 |
| Mode |  |  |
| Bolus | 24 | 22 |
| Continuous | 85 | 78 |
| Time to initiation* |  |  |
| ≤48 hours | 98 | 90 |
| >48 hours | 11 | 10 |
| Reached full enteral nutrition* |  |  |
| ≤7 days | 103 | 94 |
| >7 days | 6 | 6 |
| Complications<sup>#</sup> |  |  |
| Yes | 9 | 8 |
| No | 100 | 92 |
| Evidence of aspiration |  |  |
| Yes | 1 | 1 |
| No | 108 | 99 |

\* After NIPPV initiation
<sup>#</sup> Refer to Table S3 for details
PO = per os (by mouth), NG = nasogastric, NJ = nasojejunal, GT/JT = gastrostomy tube/jejunostomy tube

### 3.5 Physiometric Parameters and Enteral Nutrition

Finally, we examined the effect of initiating feeds on physiometric parameters, i.e. heart rate and respiratory rate. Within the same cohort of non-intubated EN patients (*n*=109), we found a significant decrease in both heart and respiratory rate after feeds were initiated (**Figure 2**). More specifically, average heart rate declined from 140 to 129 beats/min post-feeds initiation (*p* <0.001), while average respiratory rate declined from 52 to 45 beats/min post-feeds initiation (*p* <0.001). Of note, average sedation needs (dexmedetomidine and lorazepam) where not different between NPO and EN groups (**Figure S1**).

**Figure 2.**
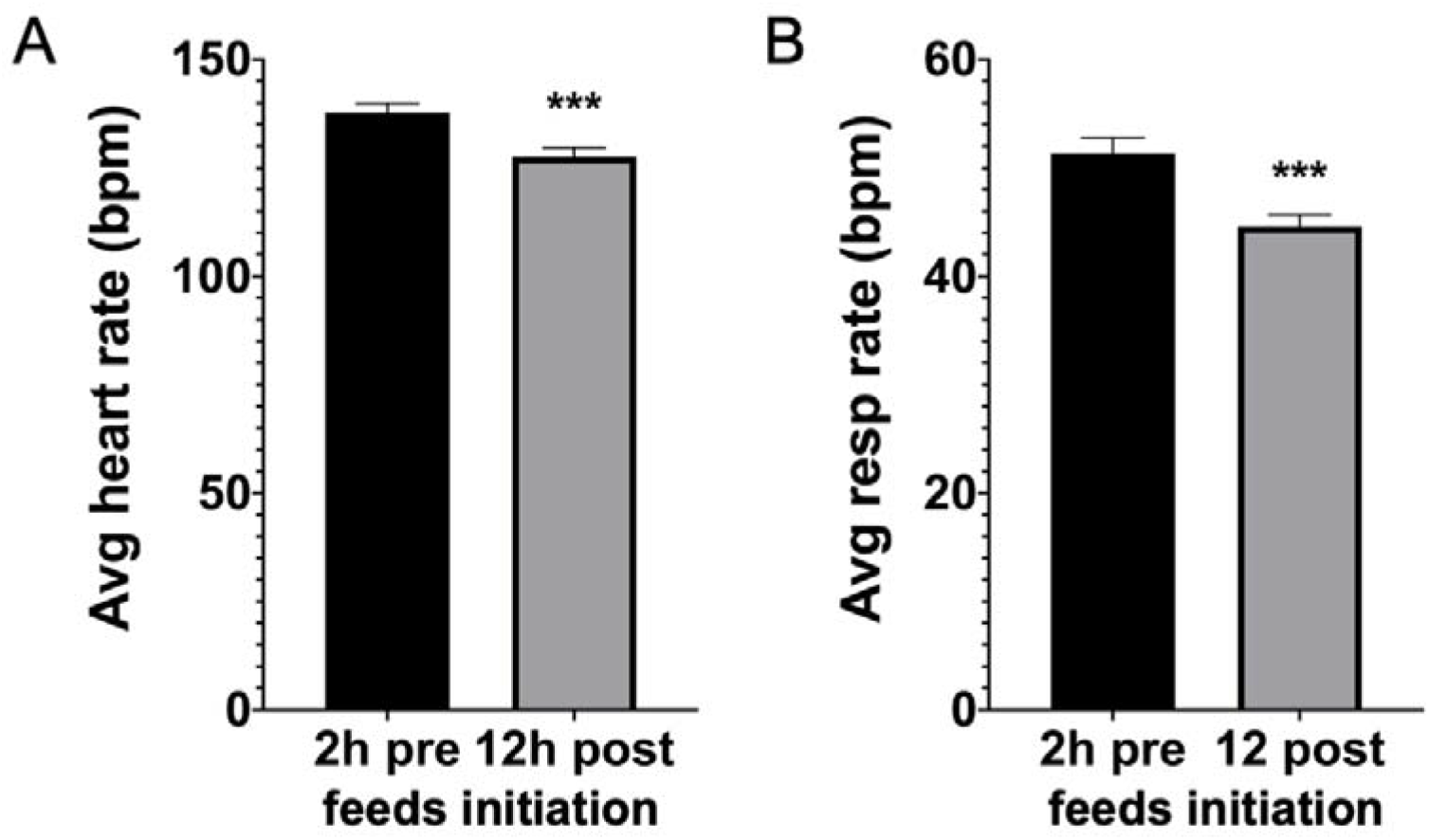
Physiometric parameters for the enteral nutrition group demonstrating (A) heart rate and (B) respiratory rate trends prior to (2h pre) and after (12h post) feeding initiation. Data presented as mean ± standard error of mean. * *p* <0.05, ** *p* <0.01, *** *p* <0.001 by Mann-Whitney U Test. Avg = average, bpm = beats/breaths per minute, resp = respiratory.

## 4. Discussion

In this retrospective single center study, we reviewed feeding practices and complications of 178 patients with critical bronchiolitis requiring non-invasive respiratory support. We found that in our institution, a majority of these patients (64%) received enteral nutrition within 48 hours of initiation of NRS, with most reaching full feeds in less than a week. Using time of NRS initiation as a surrogate for PICU transfer/admission, our practices closely align with current ASPEN[22] and ESPNIC[23] recommendations and contrast prior studies associating non-invasive support with delays in feeds initiation.[13] The overall complication rate in the EN group was relatively low with a <1% incidence of aspiration. However, none of these complications resulted in escalation of respiratory support or worsening respiratory failure. In addition to suggesting overall safety of early enteral nutrition while on NRS, physiometric data in this group associated benefit to feeding as evidenced by both lower heart rate and respiratory rate, despite no change to overall sedation needs. While this may imply improved patient comfort with feeds alone, given the retrospective nature of these data, causality cannot be discerned at this time.

We found that many demographic and clinical parameters between groups remained similar, including gender, prior history of prematurity, neurologic or genetic abnormalities, mode of NRS, pathogen number and type, illness severity, and overall mortality. That said, a key feature difference between groups was age, with patients in the EN group considerably younger with a median age of 3 months (and in turn smaller). Could this have contributed to increased provider concern regarding the risk of caloric and protein deficits in these patients, and in turn, a higher likelihood to initiate enteral nutrition in this group? Similarly, while we were surprised with the longer duration of NRS and consequently longer PICU length of stay in this group, a plausible explanation might rest in provider preference to wean NRS slower in younger patients with acute respiratory failure. Since our study was not designed to answer these questions, these observations remain a distinguishing feature of these groups that require further investigation. Notably, the incidence of intubation was lower in the EN group despite a higher incidence of bacterial superinfection. While interesting, given the low overall incidence of intubation, we cannot draw conclusions on the mitigating effects of EN on intubation rates without prospective larger scale trials.

The NPO group experienced shorter NRS duration, a higher incidence of intubation, and shorter PICU length of stay. We perceive these differences to highlight two distinct populations of patients, i.e. one relatively healthy with rapid recovery and one sicker cohort. The intubated NPO patients with a shorter duration of preceding NRS would constitute the latter, albeit smaller group, i.e. these patients were sicker at the outset. At present, we cannot determine if NPO status contributed to worsening respiratory failure in these patients secondary to agitation or caloric deficits or if patients were kept NPO appropriately due to concern for declining status. The second “healthier” group remained not only unintubated but also had a shorter duration on NRS and thus a shorter PICU length of stay (by ∼1.5 days) compared to EN group. Given the relatively older demographic of this group, this observation would not be surprising. The example of an overall healthy 10-month old female with acute respiratory failure due to bronchiolitis spending the median 48 hours in the PICU, recovering and having feeds initiated is not far-fetched. Providers might argue that an older child might tolerate NPO status better than a younger one. Further prospective studies are needed to elucidate the underlying differences between the NPO and EN groups that we have preliminarily uncovered here.

The primary limitation of our study stems from its retrospective, single-center nature. Our study showed no difference in mortality or VFD’s between groups and reduced intubation rates in the EN group. However, conclusions on the effects of early enteral nutrition on NRS duration, intubation rates, and PICU length of stay cannot be made by our investigation at this time. Pediatric studies have shown benefits of early nutrition, in both mechanically ventilated and non-invasively supported patients.[14, 24] In fact, a recently published retrospective pediatric study in 4 European PICUs reached similar conclusions to ours, reporting safety and tolerance of early enteral nutrition in patients managed on NRS.[18]

With new emphasis on the benefits of early enteral nutrition spotlighted by national societies like ASPEN and ESPNIC, the trend towards this practice noted in our study is encouraging. Aside from our study’s findings being supported, they add valuable insight on the moving practice of early enteral nutrition in critically-ill pediatric patients. As a result, we feel confident purporting not only the safety of feeding patients on NRS, but also its possible beneficial effects on patient comfort. We continue to need larger scale prospective studies specifically designed to examine the effects of early enteral nutrition on pediatric mortality, intubation rates, organ failure-free-days, and overall length of hospitalization.

## Data Availability

Data Availability Statement: Data available upon request.

## Supplementary Materials

The following are available online at www.mdpi.com/xxx/s1, Figure S1: Sedation data for the NPO and EN groups, Table S1: Demographic and clinical differences between groups (intubated only, *n* = 14), Table S2: Infecting Pathogen Characteristics (all groups, *n* = 178), Table S3: Feeding complications encountered during feeding initiation (EN group only, *n* = 9)

## Author Contributions

Conceptualization, S.M.; Data curation, A.L., V.R., N.N., J.L., J.L., and S.M.; Formal analysis, J.L., M.K., and S.M.; Investigation, A.L., V.R., N.N., J.L., and S.M; Methodology, S.M.; Supervision, S.M.; Validation, S.M.; Visualization, S.M.; Writing – original draft, A.L., V.R., and M.K.; Writing – review & editing, A.L., V.R., N.N., J.L., J.L., M.K., and S.M. All authors have read and agreed to the published version of the manuscript.

## Institutional Review Board Statement

The study was conducted according to the guidelines of the Declaration of Helsinki, and approved by the Institutional Review Board of University of Nebraska Medical Center (IRB#655-18-EP and 11/07/2017).

## Informed Consent Statement

Patient consent was waived due to the minimal risk nature of this study.

## Data Availability Statement

Data available upon request.

## Acknowledgments

We would like to acknowledge Lucinda Kustka for her assistance with preparing the IRB application for this project; and Amber Marquiss and Teresa Tobin for their support with data collection from the electronic medical record system.

## References

1. Shay, D.K., et al., Bronchiolitis-associated hospitalizations among US children, 1980-1996. JAMA, 1999. 282(15): p. 1440–6.

2. Yorita, K.L., et al., Infectious disease hospitalizations among infants in the United States. Pediatrics, 2008. 121(2): p. 244–52.

3. Wright, M., C.J. Mullett, and G. Piedimonte, Pharmacological management of acute bronchiolitis. Ther Clin Risk Manag, 2008. 4(5): p. 895–903.

4. Bont, L., et al., Defining the Epidemiology and Burden of Severe Respiratory Syncytial Virus Infection Among Infants and Children in Western Countries. Infect Dis Ther, 2016. 5(3): p. 271–98.

5. Gupta, P., B.W. Beam, and M. Rettiganti, Temporal Trends of Respiratory Syncytial Virus-Associated Hospital and ICU Admissions Across the United States. Pediatr Crit Care Med, 2016. 17(8): p. e343–51.

6. Wang, E.E.L., B.J. Law, and D. Stephens, Pediatric Investigators Collaborative Network on Infections in Canada (PICNIC) prospective study of risk factors and outcomes in patients hospitalized with respiratory syncytial viral lower respiratory tract infection. The Journal of Pediatrics, 1995. 126(2): p. 212–219.

7. Javouhey, E., et al., Non-invasive ventilation as primary ventilatory support for infants with severe bronchiolitis. Intensive Care Med, 2008. 34(9): p. 1608–14.

8. Lazner, M.R., A.P. Basu, and H. Klonin, Non-invasive ventilation for severe bronchiolitis: analysis and evidence. Pediatr Pulmonol, 2012. 47(9): p. 909–16.

9. Meduri, G.U., et al., Noninvasive positive pressure ventilation via face mask. First-line intervention in patients with acute hypercapnic and hypoxemic respiratory failure. Chest, 1996. 109(1): p. 179–93.

10. Morris, J.V., et al., Outcomes for Children Receiving Noninvasive Ventilation as the First-Line Mode of Mechanical Ventilation at Intensive Care Admission: A Propensity Score-Matched Cohort Study. Crit Care Med, 2017. 45(6): p. 1045–1053.

11. Mehta, N.M., et al., Nutritional practices and their relationship to clinical outcomes in critically ill children--an international multicenter cohort study*. Crit Care Med, 2012. 40(7): p. 2204–11.

12. Ng, G.Y.H., et al., Nutritional status, intake, and outcomes in critically ill children with bronchiolitis. Pediatr Pulmonol, 2020. 55(5): p. 1199–1206.

13. Canarie, M.F., et al., Risk Factors for Delayed Enteral Nutrition in Critically Ill Children. Pediatr Crit Care Med, 2015. 16(8): p. e283–9.

14. Leroue, M.K., et al., Enteral Nutrition Practices in Critically Ill Children Requiring Noninvasive Positive Pressure Ventilation. Pediatr Crit Care Med, 2017. 18(12): p. 1093–1098.

15. Casaer, M.P. and G. Van den Berghe, Nutrition in the acute phase of critical illness. N Engl J Med, 2014. (25): p. 2450–1.

16. Hoffer, L.J. and B.R. Bistrian, Nutrition in critical illness: a current conundrum. F1000Res, 2016. 5: p. 2531.

17. Singer, P. and S. Rattanachaiwong, To eat or to breathe? The answer is both! Nutritional management during noninvasive ventilation. Crit Care, 2018. 22(1): p. 27.

18. Tume, L.N., et al., Enteral Feeding of Children on Noninvasive Respiratory Support: A Four-Center European Study. Pediatr Crit Care Med, 2021. 22(3): p. e192–e202.

19. Buck, M.L., Dexmedetomidine use in pediatric intensive care and procedural sedation. J Pediatr Pharmacol Ther., 2010. 15(1): p. 17–29.

20. Carroll, C.L., et al., Use of dexmedetomidine for sedation of children hospitalized in the intensive care unit. J Hosp Med, 2008. 3(2): p. 142–7.

21. Yehya, N., et al., Reappraisal of Ventilator-Free Days in Critical Care Research. Am J Respir Crit Care Med, 2019. 200(7): p. 828–836.

22. Mehta, N.M., et al., Guidelines for the Provision and Assessment of Nutrition Support Therapy in the Pediatric Critically Ill Patient: Society of Critical Care Medicine and American Society for Parenteral and Enteral Nutrition. JPEN J Parenter Enteral Nutr, 2017. 41(5): p. 706–742.

23. Tume, L.N., et al., Nutritional support for children during critical illness: European Society of Pediatric and Neonatal Intensive Care (ESPNIC) metabolism, endocrine and nutrition section position statement and clinical recommendations. Intensive Care Med, 2020. 46(3): p. 411–425.

24. Srinivasan, V., et al., Early Enteral Nutrition Is Associated With Improved Clinical Outcomes in Critically Ill Children: A Secondary Analysis of Nutrition Support in the Heart and Lung Failure-Pediatric Insulin Titration Trial. Pediatr Crit Care Med, 2020. 21(3): p. 213–221.

